# Auditory beat stimulation and behavioural variant of frontotemporal dementia: a case study

**DOI:** 10.1101/2023.06.23.23290111

**Authors:** Leila Chaieb, Pawel Tacik, Michael Heneka, Juergen Fell

## Abstract

A growing number of studies suggest that auditory beat stimulation may be helpful in providing relief from symptoms of anxiety. However, evidence for this effect in clinical populations remains sparse. In this case study, we examined the effects of theta frequency monaural beat stimulation on anxiety levels and mind wandering of four patients diagnosed with behavioural variant of frontotemporal dementia (bvFTD). Over the period of a fortnight, consisting of two one-week blocks, patients were exposed daily to monaural beats at 8Hz and a pure tone control condition, in a randomized order. To assess levels of anxiety, mind wandering and measures of general well-being, patients completed the State-Trait Anxiety Inventory, Beck Depression Inventory, and Mind Wandering Questionnaire at the beginning and end of each week, as well as the Rogers’ Happy/ Sad Face Scale, before and after each stimulation. The latter consisted of subscales for anxiety, mood and pain. Scores from the questionnaires and scales pertaining to anxiety, depression and well-being indicated mixed effects of the monaural beat stimulation. A trend towards an overall decrease in mind wandering was observed across the four patients for the monaural beat stimulation condition, when compared to the control tone. These data suggest that such adjunct approaches to current therapies for anxiety are indeed possible, in patient populations.

## Introduction

A growing number of studies are investigating the role of auditory beat stimulation as a tool to modulate anxiety (Isik et al., 2017; Menziletoglu et al., 2021; Ölçücü et al., 2021; for a review see Garcia-Argibay et al., 2018). The presentation of a monaural beat, i.e. an amplitude-modulated signal given by the simple superposition of two sinusoidal tones with adjacent frequencies, offers an easy-to-apply, minimal-risk adjunct to current therapies. Research has already indicated that auditory beat stimulation may be a novel means by which to modulate cognition and mood states (for a review see Chaieb et al., 2015).

In this case study, we sought to examine the impact of monaural beat stimulation on anxiety symptoms of patients diagnosed with behavioural variant of frontotemporal dementia (bvFTD). In a recent study, monaural beat stimulation was shown to reduce levels of state anxiety in healthy individuals (Chaieb et al., 2017), and emerging research indicates that auditory beat stimulation, most prominently in the theta and alpha range, can affect changes in mood, in particular with regard to anxiety (e.g. Le Scouarnec et al., 2001; Menziletoglu et al., 2021; Weiland et al., 2011).

BvFTD is the most frequent phenotype of frontotemporal dementia (FTD), and usually occurs before the age of 65 years (Piguet et al., 2017; Rabinovici & Miller, 2010). FTDs account for approximately 10% of all dementia cases (Rosness et al., 2016), and demonstrate a diverse and heterogeneous clinical profile rendering early diagnosis complex (Cerami & Cappa, 2013; Piguet et al., 2011). FTDs result from progressive degeneration and atrophy of the frontal cortices and anterior portions of the temporal lobes. BvFTD is broadly characterised by gradual changes in behaviour and personality that reflect the progressive dissolution of neural circuits that control interpersonal conduct, emotion regulation, motivation and decision-making processes (for a review see Piguet et al., 2011). Studies vary in their estimate of the prevalence of anxiety disorders in patients with FTD. Neary et al. (1998) place it at around 6.5%, while an earlier study by Ferran and colleagues suggest it’s as high as 22% (Ferran et al., 1996; for a review see Kuring et al., 2018). Anxiety levels can be elevated further when patients, who can become highly dependent upon their caregivers, feel that they are not within the sight of their caregiver (Caselli & Yaari, 2007).

Due to the complex symptomatology and progressive nature of bvFTD, a therapeutic option that is easy to implement, and that can effectively modulate anxiety levels in patients without additional pharmacological agents would be useful. Indeed, some studies have applied music therapy with the goal of alleviating anxiety symptoms in patients with FTD (Langhammer et al., 2019; Raglio et al., 2012; Spinosa et al., 2022). This form of therapy has been shown to be effective in treating anxiety symptoms, but requires careful consideration of the material used, and ideally collaboration with a music therapist (Raglio et al., 2012). In this case, monaural beat stimulation may offer a simpler means of reducing anxiety symptoms in bvFTD patients.

To this end, we invited patients diagnosed with bvFTD to participate in the current study. They were asked to listen to either a 20-minute monaural beat sound file (modulation frequency: 8 Hz; carrier frequency: 220 Hz), or a sine wave control sound file (frequency: 220 Hz), once a day over six days, and then the alternate recording, again over a six-day period (one day pause between stimulation periods). Patients were also asked to complete a number of questionnaires and scales related to anxiety (State-Trait Anxiety Inventory), depression (Beck Depression Inventory), mind wandering (Mind Wandering Questionnaire), and a visual numeric scale with items related to anxiety, negative mood and pain (Rogers’ Happy/ Sad Face Scale). Since bvFTD patients experience profound cognitive, personality and behavioural changes that affect well-being, we sought to examine whether monaural beat stimulation may, besides anxiety, also impact upon these aspects of disease progression by including the Beck Depression Inventory, as well as the Mind Wandering Questionnaire.

Based on the outcomes from previous studies, we hypothesised reductions in anxiety scores based on the STAI, as well as in momentary anxiety levels based on the daily visual-numeric scale for the 8 Hz monaural beat condition, compared to the sine wave control stimulation. Furthermore, we hypothesised a decrease in mind wandering for monaural beats versus control stimulation, similar to the outcomes that we have observed in healthy participants (Chaieb et al., 2022).

## Methods

### Patients

A brief clinical description of the four patients, which completed the study, is provided in the following section.

Patient 1 had a 2 year history of progressive behavioural impairment including symptoms of motor neuron disease that initially affected the lower limbs and that later also affected the arms. Upon clinical neurological examination, the patient showed symptoms of upper motor predominant amyotrophic lateral sclerosis (ALS) with probable behavioural variant frontotemporal dementia (bvFTD), in accordance with the international consensus criteria for diagnosing bvFTD (Rascovsky et al., 2011). An MRI scan showed mild general brain atrophy, most pronounced in the anterior region of the right temporal lobe. A cognitive examination in the form of the extended version of the Consortium to Establish a Registry for Alzheimer’s disease (CERAD plus) test battery, revealed mild cognitive impairment of multiple domains. Analysis of cerebrospinal fluid (CSF) biomarkers showed an elevated level of phosphorylated neurofilament heavy-chain (pNfh) and a decreased level of amyloid beta (1-42), whereas total tau (ttau), phosphorylated tau (ptau), amyloid beta 42/40 ratio, and amyloid beta 42/ptau ratio were normal. A fiber optic endoscopic evaluation of swallowing (FEES) test revealed a level 3 dysphagia. The patient was placed on an oral suspension of riluzole.

Patient 2 had a 3 year history of progressive behavioural impairment. The patient was diagnosed with probable bvFTD. An MRI revealed severe frontotemporal atrophy. The patient’s cognitive profile, examined using the CERAD plus test battery, was consistent with the stage of moderate dementia. Clinical neurological examination, as well as CSF biomarker analysis did not show any relevant abnormalities. The patient also presented symptoms of apathy, and so sertraline treatment was prescribed at a dose of 50 mg/day.

Patient 3 was diagnosed with bvFTD due to a p.P301L mutation in microtubule-associated protein tau (MAPT) gene. At around age 50-55 years, the patient began complaining about progressive behavioural abnormalities with executive dysfunction. Cerebral MRI scans over a duration of the last 5 years showed a progressive volume reduction accentuated in the frontotemporal lobes. A clinical neurological examination and a CSF biomarker analysis were normal. Cognitive deficits in the CERAD plus test battery reached the level of multiple-domain mild cognitive impairment.

Patient 4 was diagnosed with both FTD and ALS. One year prior to the established diagnosis, the patient had started developing motor and behavioural symptoms. Due to a muscle weakness, the patient’s gait had become unsteady, and they showed impairment in fine motor skills. The patient’s family observed that their behaviour had become impulsive, and was characterised by rash actions accompanied by reduced empathy. Upon clinical neurological examination, symptoms of impairment of both the upper and the lower motor neurons were identified in three body regions. A CSF analysis showed elevated pNfh and total tau levels. In contrast, the diagnostic MRI scan did not reveal anything contributory. The patient’s behavioural symptoms met the criteria of probable bvFTD according to the criteria of Rascovsky and al. (2011). Upon neurological examination, a mild impairment of executive function, speech and memory was detected. A FEES test revealed mild dysphagia. The patient was then prescribed riluzole.

### Study procedure

11 patients diagnosed with probable bvFTD according to Rascovsky’s criteria (Rascovsky et al., 2011) were recruited into the study by a consultant neurologist at the University Hospital Bonn (author PT). The patient and accompanying caregiver were informed of the details of the study during a clinical consultation with the consultant neurologist. A step-by-step handbook (including consent forms and documentation required by the Ethics Committee at the University of Bonn), was given to each of the patients along with an MP3 player (together with in-ear headphones) containing the two sound files; an 8 Hz monaural beat recording and a sine wave control tone recording. During the consultation, patients and caregivers were fully informed of the requirements of the study, which included a detailed explanation using the handbook as a guide. They were then instructed on how to administer the auditory beat stimulation using the portable MP3 player. All details discussed during the initial patient meeting could also be found in the study handbook, for later reference for the patients and caregivers. Written, informed consent was given either by the patient or a certified responsible party (relative or caregiver), and obtained during the initial consultation. The study was conducted in accordance with the Declaration of Helsinki, and all experimental procedures/ questionnaires were approved by the Ethics Committee of the Medical Faculty of the University of Bonn.

The duration of the study was two weeks, and comprised of two seven-day blocks with day seven being designated a rest day. At the beginning of each week, patients were asked to complete the State-Trait Anxiety Scale (STAI), the Beck Depression Inventory BDI-II), and the Mind Wandering Questionnaire (MWQ). Further to this, patients were asked to assess twice daily their level of anxiety, mood and pain using the Rogers’ Happy/ Sad Face Scale. They did so prior to and after stimulation (see Figure 1).

**Figure 1:**
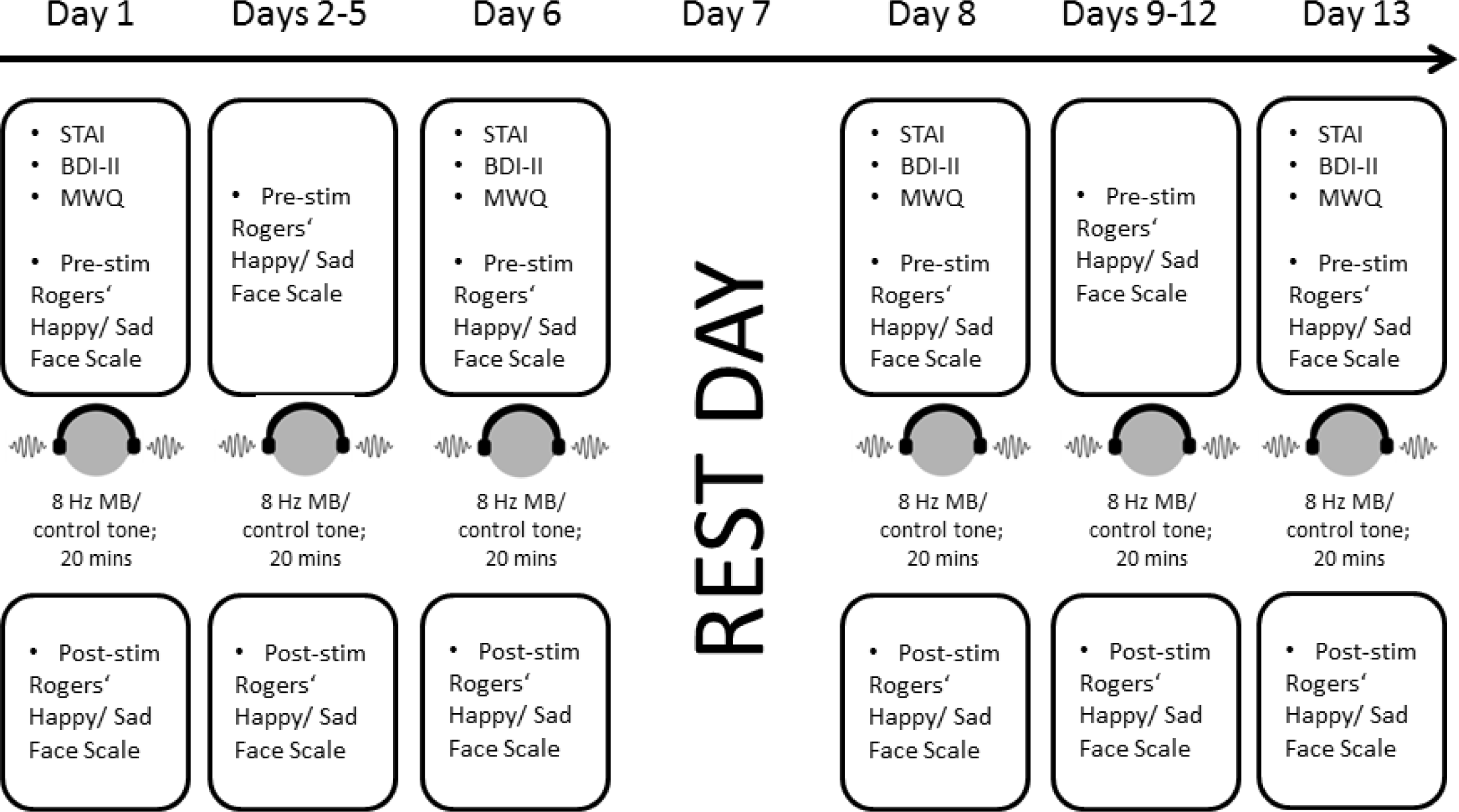
An overview of the study. Patients recruited into the study were asked to complete the STAI, BDI-II, MWQ and pre- and poststimulation visual numeric scales (Rogers’ Happy/ Sad Face Scale). The study was conducted over two one-week blocks during which patients received a daily session of auditory beat stimulation for 20 mins.

The order of auditory stimulation conditions across patients was randomised and counterbalanced. Patients and caregivers were blinded as to which stimulation condition they received or administered, and in which order. A summary of the order of auditory beat stimulation for the patients that completed the study, as per each week, is as follows: patient 1: control tone/ monaural beat; patient 2: monaural beat / control tone; patient 3: monaural beat/ control tone; patient 4: control tone/ monaural beat.

As a result of the demands of the study (the number of questionnaires required both weekly and daily, administering the daily course of auditory stimulation, returning handbooks and questionnaires), a significant number of patients were unable to complete the study (N= 7). These individuals were not included in the final descriptive analysis. Of the 4 remaining patients, four completed the MWQ, two the BDI, one the STAI, and four the Rogers’ Happy/ Sad Face Scales.

### Neuropsychological Scales and Questionnaires

#### The State-Trait Anxiety Inventory (STAI)

The STAI Inventory was used to assess the patients’ state (STAI-S) and trait (STAI-T) anxiety. The two subscores differentiate between transient feelings of anxiety (state anxiety), i.e. those related to a particular context like an event, and anxiety levels that are more of a personal characteristic of an individual (trait anxiety: Spielberger et al., 1983). Patients responded to 20 statements contained in each subscore, using a 4-point Likert scale (not at all: 1; a little: 2; quite: 3; very: 4). The STAI Inventory was completed at the beginning and end of each week.

#### The Beck Depression Inventory (BDI-II)

The BDI is a 21-item multiple-choice questionnaire (Beck et al., 1996). The BDI was used to assess symptoms of depression, and was administered at the beginning and end of each week, prior to the six-day daily course of auditory beat stimulation. The BDI is categorised into the following scoring ranges: 0-13 minimal; 14-19 mild; 20-28 moderate and 29-63 severe.

#### The Mind Wandering Questionnaire (MWQ)

The Mind Wandering Questionnaire is a short-form scale intended to measure the propensity to mind wander (Mrazek et al., 2013). The questionnaire consists of five items that evaluate trait levels of mind wandering. Trait mind wandering is scored across a 6-point Likert scale (1: almost never; 2: very infrequently; 3: somewhat infrequently; 4: somewhat frequently; 5: very frequently; 6: almost always). Patients completed this questionnaire, also at the beginning and end of each week.

#### The Rogers’ Happy/ Sad Face Scale

The Rogers’ Happy/ Sad Face Scale is a visual-numeric scale that was intended to capture the momentary subjective levels of anxiety, mood and pain. Patients had to grade their feelings according to a corresponding face icon which represented a ‘happy face’ at the most positive end (value of 0) of the graded scale, all the way to a ‘crying face’ icon at the most negative end of the scale (value of 4). Patients completed this scale twice daily, once immediately prior to the auditory stimulation, and then immediately after the stimulation had concluded.

#### Auditory beat stimulation

To examine potential effects of monaural beat stimulation, two auditory recordings (MP3 files) were generated: a monaural beat (modulation frequency: 8 Hz; carrier frequency: 220 Hz) and a sine wave control tone (frequency: 220 Hz). The monaural beat was constructed in the following way: two sine waves with frequencies of 216 and 224 Hz were superposed on each other, resulting in an amplitude modulation of 8 Hz. The control tone, however, was a pure sine wave at 220 Hz. Thus, by construction, the frequency of the control tone was identical to the carrier frequency of the monaural beat. The auditory recordings were created using Tone Generator (NCH software, Canberra, Australia). Both auditory stimulation recordings were played using in-ear headphones with a preset sound pressure level (SPL) of 70 dB. Each recording lasted for 20 minutes.

#### Data and statistical analyses

Data was compiled from the patients’ responses to the questionnaires and scales included in the patient handbook. Based on these, we analysed increases or decreases in post-versus pre-stimulation scores according to auditory stimulation condition, i.e. the 8 Hz monaural beat or the 220 Hz sine wave control tone. The three questionnaires were filled out biweekly (BDI-II, STAI, and the MWQ). The Rogers’ Happy/ Sad Face Scale was completed twice daily, before and after each stimulation session. Due to a high number of dropouts (7/11 patients), data from the biweekly questionnaires and the Rogers’ Happy/Sad Face Scale was examined on an individual patient basis. For the MWQ (data from 4 patients), we additionally evaluated the group results for the two stimulation conditions. For the Rogers’ Happy/ Sad Face Scale we compared score differences (post-versus pre-stimulation) across days between stimulation conditions. Statistical comparisons between stimulation conditions were performed using Wilcoxon signed rank tests. These tests were conducted one-sided for the anxiety and mind wandering measures, for which we had specific hypotheses (see Introduction), and two-sided otherwise.

## Results

### The State-Trait Anxiety Inventory (STAI)

Only one patient was able to complete the STAI Inventory. Patient 3 showed a decrease in the state-anxiety subscore for the monaural beat stimulation condition (pre: 55/ post: 38), and an increase for the control tone (pre: 41/ post: 49). Also for the trait-anxiety subscore, patient 3 showed a decrease for the monaural beat condition (pre: 45/ post: 41), and an increase for the control tone (pre: 37/ post: 44).

### The Beck Depression Inventory (BDI-II)

Only two patients completed this questionnaire sufficiently for analysis. Patient 1 showed a decrease in BDI scores for both the monaural beat condition (pre: 21/ post: 14) and the control condition (pre: 15/ post: 12). Patient 3 showed a decline in score for the monaural beat condition (pre: 17/ post: 8); whilst an increase in BDI score for the control condition (pre: 6/ post: 8).

### The Mind Wandering Questionnaire (MWQ)

For the MWQ, four patients were able to fill out the short form questionnaire. Regarding the individual sum scores, patient 1 displayed a slight increase for the beat stimulation condition (pre: 17/ post: 18) and a slight decrease for the control tone (pre: 18/ post: 17). Patient 2 had a pronounced decrease in MWQ score for the monaural beat condition (pre: 13/ post: 7), and an increase for the control condition (pre: 7/ post: 9). Patient 3 showed a similar pattern; a strongly decreased score for the beat condition (pre: 15/ post: 7) and an increase for the sine wave control tone (pre: 9/ post: 11). Patient 4, however, showed an increase in MWQ score for the monaural beat stimulation condition (pre: 5/ post: 8), and a pronounced increase for the control tone (pre: 7/ post: 15).

Furthermore, we evaluated the difference between the post- and prestimulation scores for each of the 5 questionnaire items and the sum score, across all patients. For the sum score and all item scores except item 4, trends towards decreases of the mind wandering differences (after versus prior to) for monaural beat stimulation compared to the control tone were found (Wilcoxon signed rank tests (one-sided); item 1: p = 0.071, z = 1.473; item 2: p = 0.099, z = 1.289; item 3: p = 0.098, z = 1.3; item 5: p = 0.09, z = 1.342; sum score: p = 0.072, z = 1.461).

### The Rogers’ Happy/ Sad Face Scale

For the visual-numeric scale, for each patient the (pre- and poststimulation) sum scores for each of 3 items (anxiety, negative mood, and pain) across the six-day study period, for the monaural beat stimulation and control tone condition, were calculated.

For the anxiety ratings, patient 1 showed an increase in anxiety scores for the monaural beat condition (pre: 1.0/ post: 1.17) and no change for the control tone (pre: 0.5/ post: 0.5). Patient 2 demonstrated no change in anxiety score for the beat stimulation condition (pre: 2.0/ post: 2.0), nor for control condition (pre: 2.0/ post: 2.0). Patient 3 showed a decrease in anxiety score for the beat condition (pre: 0.33/ post: 0), and no change in the control condition (pre: 0/ post: 0). Patient 4 showed no change in anxiety rating for the monaural beat condition (pre: 0/ post: 0), but an increase for the control tone (pre: 0/ 1.0). Across days, for this patient a trend was found for a decrease of anxiety score differences (post versus pre) for monaural beats compared to control stimulation (Wilcoxon signed rank test (one-sided): p = 0.079, z = 1.414).

For the negative mood item, patient 1 showed an increase for the monaural beat condition (pre: 1.17/ post: 1.33), and a decrease for the control tone (pre: 1.33/ post: 1.17). Patient 2 showed no change in the beat condition (pre: 1.67/ post: 1.67), but a decrease for the control condition (pre: 2.0/ post: 1.6). Patient 3, a decline in negative mood for the monaural beat condition (pre: 1.17/ post: 0.17), and also a decline for the control tone (pre: 1.33/ post: 0). Patient 4, however, showed an increase in negative mood scores for the monaural beat (pre: 0.2/ post: 0.4), but also an increase for the control tone (pre: 1.83/ post: 2.5).

For the pain item on the rating scale, patient 1 demonstrated an increase in the monaural beat condition (pre: 0.6/ post: 1.0), but also an increase for the control tone (pre: 0.17/ post: 0.33). Patient 2 showed no change for the beat condition (pre: 1.83/ post: 1.83), and a decrease in the control condition (pre: 2.2/ post: 2.0). Similarly, patient 3 demonstrated no change for the beat condition (pre: 0/ post: 0), and also indicated no change for the control condition (pre: 0/ post: 0). Patient 4 showed no change after monaural beat stimulation for the pain item (pre: 0/ post: 0), but a decline for the control tone (pre: 0.83/ post: 0.67).

## Discussion

In this case study we aimed to investigate the impact of 8 Hz monaural beat stimulation on the anxiety levels of patients diagnosed with bvFTD. In addition, we also examined mind wandering, momentary mood and pain. Daily changes in anxiety, negative mood, and pain prior to and after monaural beat stimulation, compared to the sine wave control tone, were measured using the Rogers’ Happy/ Sad Face Scale (a visual numeric scale). As a major limitation, a large number of patients did not complete the study, and we were only able to examine data of 4 patients.

With regard to our initial hypothesis related to anxiety, we were able to see a trend towards a reduction in the momentary anxiety rating differences (post-versus pre-stimulation) for monaural beats compared to control stimulation in patient 4. Moreover, a beat stimulation-related decline in both state and trait anxiety scores was found for the single patient (patient 3) who completed the STAI Inventory, while state and trait anxiety scores increased for the control condition. Also, momentary anxiety ratings decreased in this patient after beat stimulation and did not change for the control condition. An earlier study by our group has shown that monaural beats can exert a positive effect on state anxiety in healthy participants, measured using the STAI-S (Chaieb et al., 2017). Even though we were only able to collect data from a very small number of patients, such studies looking at adjunct therapies are becoming increasingly more relevant. There is still some discussion concerning the practicability of non-invasive stimulation methods for patient populations (for a review see Sanches et al., 2020). With regard to bvFTD, recent studies suggest that an anxiety disorder is a risk factor (Rasmussen et al., 2018), whereas other perspectives view anxiety as a symptom in the course of FTD, especially in the prodromal phase (Kuring et al., 2018). In a review, Levenson and colleagues state that “one challenge with FTD is determining whether behavioural symptoms that are often associated with anxiety in other patient groups are actually associated with subjective feelings of anxiety when manifest by patients with FTD” (Levenson et al., 2014). This citation indicates that patients may also experience difficulties in communicating their own subjective feelings of anxiety.

Of considerable interest was the data derived from the mind wandering questionnaire. We observed a trend for a decrease of mind wandering for the monaural beat stimulation condition, compared to the control tone. The results from a recent study also reported a reduction in mind wandering during monaural theta beat stimulation, in individuals exhibiting high trait-level mind wandering (Chaieb et al., 2022). In an earlier study, O’Callaghan and colleagues demonstrated that during a visual attention task, bvFTD patients displayed significantly reduced capacity for mind wandering, but this was interestingly compensated for by a significant increase in stimulus-bound thought (O’Callaghan et al., 2019). Such studies are important, as they highlight the current state of limited knowledge concerning the altered patterns of mind wandering in clinical populations.

The early-onset and gradual progression of bvFTD can render the symptoms of this disorder difficult to manage, even considering current therapies. This case study suggests that auditory beat stimulation, in this case monaural beat stimulation, may offer an opportunity for patients to alleviate anxiety symptoms effectively and safely without additional pharmacological agents. Furthermore, we have seen that monaural beat stimulation may also modulate mind wandering in patients diagnosed with bvFTD.

## Author contributions

(as per the CRediT author statement): Conceptualization: Juergen Fell, Michael Heneka, Pawel Tacik and Leila Chaieb; Methodology: JF, MH, PT and LC; Data curation: PT and LC; Formal analysis: JF and LC; Writing-Original draft preparation: LC, JF and PT; Investigation: PT and LC; Visualisation: JF, MH, PT and LC; Supervision: JF and MH; Validation: JF, PT and LC; Writing-review and editing: LC, JF, PT and MH.

## Data Availability

Data is available upon request. This has been stated within the manuscript.

